# A statistical algorithm for outbreak detection in a multi-site setting: the case of sick leave monitoring

**DOI:** 10.1101/2020.09.22.20199406

**Authors:** Tom Duchemin, Angela Noufaily, Mounia N. Hocine

## Abstract

Surveillance for infectious disease outbreak or for other processes should sometimes be implemented simultaneously on multiple sites to detect local events. Sick leave can be monitored accross companies to detect issues such as local outbreaks and identify companies-related issues as local spreading of infectious diseases or bad management practice. In this context, we proposed an adaptation of the Quasi-Poisson regression-based Farrington algorithm for multi-site surveillance. The proposed algorithm consists of a Negative-Binomial mixed effect regression with a new reweighting procedure to account for past outbreaks and increase sensitivity of the model. We perform a wide range simulations to assess the performance of the model in terms of False Positive Rate and Probability of Detection. We propose an application to sick leave rate in the context of COVID-19. The proposed algorithm provides good overall performance and opens up new opportunities for multi-site data surveillance.

## 1 Introduction

The increasing flow of data and the recent epidemic threats have increased the need for the development of robust epidemiological surveillance. Epidemiological surveillance is not limited to the study of new cases of a disease, but has been widely used in pharmacovigilance, to detect adverse events related to drug and vaccine consumption [1], and in syndromic surveillance to detect unusual concentration of a virus in waste water or an unusual amount of Google queries as an early signal of an outbreak [2, 3, 4]. The diversity of these data calls for new or adapted methods that could fit to different issues.

Many reviews of epidemiological surveillance methods have already been performed [5, 6, 7, 8] and many statistical techniques have been used for this purpose: regression, time series, statistical process control, spatio-temporal methods for instance [5]. Within the framework of regression models, a widely-used algorithm developed by Farrington *et al*. [9] uses a Quasi-Poisson regression adjusted for trend and seasonality and reweighted to account for past outbreaks [10]. The Farrington algorithm was extensively validated with simulations and was adapted to improve the weighting procedure and seasonality by Noufaily *et al*. [11]. Another well-known algorithm is RAMMIE [12] and uses mixed-model Poisson regression and Negative-Binomial regression. In this second algorithm, a mixed effect was included to monitor infectious diseases at local levels. Both algorithms are used routinely by health authorities [10, 12].

Our methodological development is motivated by the analysis of sick leave data. Companies are places where lots of diseases, as infectious disease or stress [13, 14], can spread. Those diseases could lead to sick leaves and then to sick leaves outbreaks if actions are not taken in time. Monitoring sick leave could help companies to identify ongoing issues and to provide a better environment for workers.

Epidemiological surveillance is already used to monitor occupational health issues such as work-related injuries to identify ongoing issues [15], to monitor school absenteeism or agregated sick leave data at the regional level and to identify influenza outbreaks [16, 17]. To our knowledge, no method was developed on the specific case of the surveillance of sick leave rate in multiple companies.

Monitoring sick leave data raised specific methodological issues: sick leave rate of companies shows a strong seasonal pattern since it is highly correlated to seasonal infectious diseases such as influenza [18] and should then be adjusted for trend and seasonality. Moreover, it should also be adjusted for other covariates since sick leave rate is associated with exogenous events as school holidays or with the population of workers in each company (age is for instance associated with sick leave rate [19]). To adjust for those covariates, a mixed effects regression model should also be fitted, with the companies being included as a random effect in the model. To solve these issues linked to sick leave data, we propose a model inspired by Farrington and RAMMIE: a Negative-Binomial mixed effects regression algorithm with reweighting procedure to account for past outbreaks. We propose to assess it with extensive simulation and present an application to sick leave data.

In this paper, we adapt the Farrington algorithm to the case of a multi-site surveillance. Section 2 describes the mixed model based algorithm. Section 3 sets out the design of the simulation to study the algorithm performance for outbreak detection and Section 4 describes the results in terms of False Positive Rate (FPR) and Probability of Detection (POD). In Section 5, we present an application to sick leave data. Finally, we discuss our findings and their implications in Section 6.

## 2 A mixed model for outbreak detection

To determine if a count outcome is unusually high and to detect outbreaks, we use the same ideas as in the Farrington algorithm [9]. The Farrington algorithm output is an outbreak threshold based on a Quasi-Poisson regression and reweighted to downweight previous outbreaks. Our algorithm uses these ideas and adapts them to a mixed model context.

### 2.1 The developed algorithm

To determine if *Y*_*i,T*_, the count outcome in a site *i* ∈ {1, …, *N*} with *N* > 0 at week *T* > 0 is an outbreak, we proceed in three steps.

**First step** First, we fit a Negative-Binomial regression on the past counts *Y*_*i,t*_ with *t* ∈ {0, …, *T*} adjusting for trend (*η*), seasonality (*γ*), covariates (*β*) and a random effect representing the sites’ effects *u*_*i*_ ∼ *N* (0, *σ*^2^):

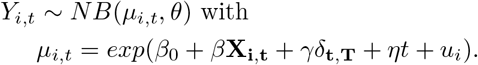

*μ*_*i,t*_ is the mean of the Negative Binomial distribution and *θ* is the dispersion parameter such that 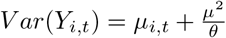. **X**_**i,t**_ is a matrix of covariates and *β* are the associated coefficients.

*δ*_*t,T*_ is a matrix of level factors adjusting for seasonality similarly to the flexible Farrington algorithm [11] to give more weights to the comparable periods in past years. Each column of *δ*_*t,T*_ describes a period: a first reference 7-week period (corresponding to weeks *T* ± 3 weeks) and nine 5-week periods in each year. *γ* are the associated parameters. It is thus a matrix with 10 columns (one per period) which gives 1 for the period associated with t and 0 otherwise.

To avoid adaptation of the model to emerging outbreaks, we exclude the 26 most recent weeks from the baseline data and we only fit the regression on the previous weeks.

**Second step** In a second step, we reweight the outliers of the training dataset to underweight past alerts and to fit a more robust outbreak threshold. We use the following weight function. ∀*i, t* > 0,

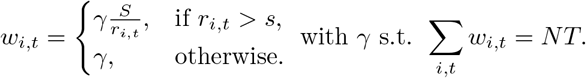

*S* > 0 is a constant controlling for the strictness of the reweighting and *r*_*i*_ are the Pearson residuals of the model and are defined as:

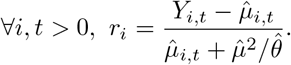

**Third step** The third step is the computation of the outbreak threshold. We fit a new Negative-Binomial regression with the previous reweighting to give less importance to past outbreaks. The outbreak threshold of company *i* ∈ {1, …*N*} is defined as 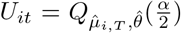 with 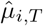 and 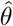 the respective estimates of *μ*_*i,T*_ and *θ* retrieved from the second regression, 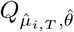, the quantile function of a Negative Binomial distribution with parameters 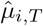 and 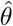 and 1 − *α* the chosen confidence level.

All the sites could be hierarchized and a specific site is flagged for further investigation if the outbreak threshold *U*_*it*_ is exceeded. An exceedance score can be defined as:

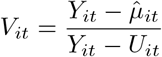

All the analyses are performed in R and with the help of the package *glmmTMB* [20]. Codes used in this article can be read from an online deposit referenced in the **Supporting Information**.

### 2.2 Comparaison with Farrington and Farrington flexible algorithms

Our model provides an adaptation of the Farrington algorithm in the context of multi-site data. The main change is the inclusion of a random effect and it resulted in the modification of the algorithm.

First, we fitted a Negative-Binomial regression model instead of a Quasi-Poisson model. We used a Negative-Binomial because a quantile function was needed to derive our threshold and this function is not defined for the Quasi-Poisson model.

Second, the formula for the threshold in the original Farrington algorithm used a normal approximation thanks to Taylor expansion and to power transformation. Our threshold is more straightforward as it only used the quantiles of the estimated distribution as was also proposed in Farrington Flexible [11].

Finally, we chose a different weight function. The weight function used in the Farrington algorithm includes a hat matrix that is not straightforward in a mixed model framework. Our function is however similar since it used standardized Pearson residuals instead of standardized Anscombe residuals.

## 3 Simulation study

We will investigate the validity of the model using extensive simulation study. We will first describe the simulated datasets and their associated scenarios and we will then propose some exploratory analyses. the procedure is similar to Noufailly *et al*. [11] in order to allow for a comparison.

### 3.1 Simulated datasets

#### Baseline data

We generate data using a negative binomial model of mean *μ* and variance 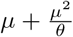 with *θ* > 0 a dispersion parameter. To be consistent with Noufaily *et al*., we reparametrize the Negative Binomial distribution such that its variance equals to *ϕμ*. We then define *θ* as: 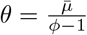 with 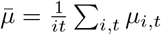

*μ*_*i,t*_ is defined for *i* ∈ {1, …, *N*} and *t* ∈ {1, …, *T*} as:

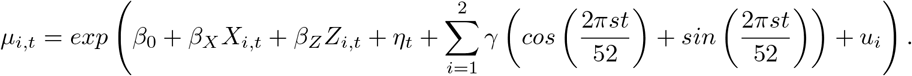

*μ*_*i,t*_ is then defined by an intercept *β*_0_, two covariates *X*_*i,t*_ and *Z*_*i,t*_ we will define later, a trend *θ*, a seasonality we defined with Fourier terms and a random effect *u*_*i*_ ∼ 𝒩 (0, *σ*^2^) with *σ* > 0.

In practice, we expect to have continuous and discrete covariates. The covariates should be quite stable in each site but can be very different from one site to another. We then simulate X and Z as the following:

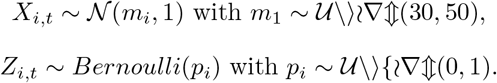

In all of the simulations, we set *N* = 50 sites and *T* = 312 weeks which correspond to 6 years of data. The most recent 52 weeks constitue the current data set we use to evaluate the model and the previous 260 weeks constitue the baseline data we will use to fit the model. We will also fix the weight threshold *s* to *s* = 2.5 and the type I error to *α* = 0.95.

#### Outbreaks

We simulated outbreaks as follows:

1. We randomly selected four weeks from the baseline data and one week from the current data;
2. For each week *t*_0_ > 0, we randomly generated the outbreak size with a Poisson random variable of mean equal to *k* > 0 times the standard deviation of the baseline count at *t*_0_;
3. We finally randomly distributed these cases in time using a lognormal distribution of mean 0 and standard deviation 0.5.

In the baseline scenarios, we will use *k* = 3 to simulate medium outbreaks. Other *k* will be chosen to test the performance of the model in different contexts.

#### Simulated scenarios

To evaluate the robustness of the model to a wide range of data sets we can meet in real life, we generate our simulations from 32 parameter combinations described in Table 1. We try different baseline volumes (given by *β*_0_), different trends and covariates (given by *η, β*_*X*_ and *β*_*Z*_), different overdispersion (given by *θ*) and different standard deviations for the random effect (given by *σ*^2^).

**Table 1:** Parameters used to generate the 32 scenarios.

| Scenario | $\beta_0$ | $\eta$ | $\beta_X$ | $\beta_Z$ | $\phi$ | $\sigma^2$ |
| --- | --- | --- | --- | --- | --- | --- |
| 1 | 1 | 0 | 0 | 0 | 1,5 | 0,5 |
| 2 | 1 | 0 | 0 | 0 | 1,5 | 1,5 |
| 3 | 1 | 0,0025 | 0 | 0 | 1,5 | 0,5 |
| 4 | 1 | 0,0025 | 0 | 0 | 1,5 | 1,5 |
| 5 | 1 | 0 | -0,5 | 1 | 1,5 | 0,5 |
| 6 | 1 | 0 | 0,5 | 1 | 1,5 | 1,5 |
| 7 | 1 | 0,0075 | -0,5 | 0,5 | 1,5 | 0,5 |
| 8 | 1 | 0,0075 | -0,5 | 0,5 | 1,5 | 1,8 |
| 9 | 3 | 0 | 0 | 0 | 1,5 | 0,5 |
| 10 | 3 | 0 | 0 | 0 | 1,5 | 2 |
| 11 | 3 | 0,0025 | 0 | 0 | 1,5 | 0,5 |
| 12 | 3 | 0,0025 | 0 | 0 | 1,5 | 2 |
| 13 | 2 | 0,0025 | -1 | 1 | 1,5 | 0,5 |
| 14 | 2 | 0,0025 | -1 | 1 | 1,5 | 1 |
| 15 | 2 | 0,0075 | -0,5 | 0,5 | 1,5 | 0,5 |
| 16 | 2 | 0,0075 | -0,5 | 0,5 | 1,5 | 1,8 |
| 17 | 1,5 | 0 | 0 | 0 | 3 | 0,5 |
| 18 | 1,5 | 0 | 0 | 0 | 3 | 1,5 |
| 19 | 1,5 | 0,0025 | 0 | 0 | 3 | 0,5 |
| 20 | 1,5 | 0,0025 | 0 | 0 | 3 | 1,5 |
| 21 | 0,5 | 0,0025 | -1,5 | 1,5 | 3 | 0,5 |
| 22 | 0,5 | 0,0025 | -1,2 | 1,2 | 3 | 1,5 |
| 23 | 0,5 | 0,0075 | -0,5 | 0,5 | 3 | 0,5 |
| 24 | 0,5 | 0,0075 | -0,5 | 0,5 | 3 | 1,5 |
| 25 | 3 | 0 | 0 | 0 | 3 | 0,5 |
| 26 | 3 | 0 | 0 | 0 | 3 | 1,5 |
| 27 | 3 | 0,0025 | 0 | 0 | 3 | 0,5 |
| 28 | 3 | 0,0025 | 0 | 0 | 3 | 1,5 |
| 29 | 3 | 0,0025 | -1,2 | 1,2 | 3 | 0,5 |
| 30 | 2 | 0,0025 | -1,2 | 1,2 | 3 | 1,5 |
| 31 | 3 | 0,0075 | -0,5 | 0,5 | 3 | 0,5 |
| 32 | 2 | 0,0075 | -0,5 | 0,5 | 3 | 1,5 |

For each scenario, we perform 5 replications of *N* = 50 companies for *T* = 52 weeks. We perform only 5 replications because the computation time of the algorithm is long as we will see latter.

### 3.2 Evaluation metrics

We evaluate the performance of the model using criteria already suggested in Noufaily *et al*. to ensure a comparison between the two models. The two criteria evaluate the performance of the model when there is an outbreak and where there is no outbreak.

For each replicate of the simulations, we first calculated the False Positive Rate (FPR) as the proportion of observations where the observed value exceeded the threshold in the absence of any current outbreak. The FPR is a rate per week.

The second criterion we calculated is the Probability of Detection (POD): it describes the probability that a true outbreak is detected. A true outbreak is detected when the observed value exceeded the threshold at least one week in the presence of a current outbreak. The POD is defined as the proportion of outbreaks detected among the 50 companies in each replicate.

For both of these criteria, we will compute it for each of the 5 replicates and we will report the average value and the minimum and maximum values across those replicates, to briefly assess the variability of these criteria between simulations.

## 4 Results of the simulation

We run our algorithm on the most recent 52 weeks of the 32 simulation scenarios in Table 1. For each scenario, we run 5 replicates from each of 50 companies. Hence, we run 416,000 simulations in total.

We first perform the simulation for a fixed medium outbreak size (*k* = 3) and reweighting threshold (*s* = 2.5). We then undertake exploratory analyses to investigate the most appropriate value for *s*.

### 4.1 False positive rates

Figure 1 shows the FPRs we obtained for *α* = 0.05, *k* = 3 and *s* = 2.5. Each point represents the median of the five simulations and the vertical line represents the range of FPRs for those 5 iterations. The nominal FPR is 0.025 and we see that the actual FPRs are a bit lower. Scenarios with no trend and with low random effect presents higher FPR but still lower than 0.025 in median.

**Figure 1:**
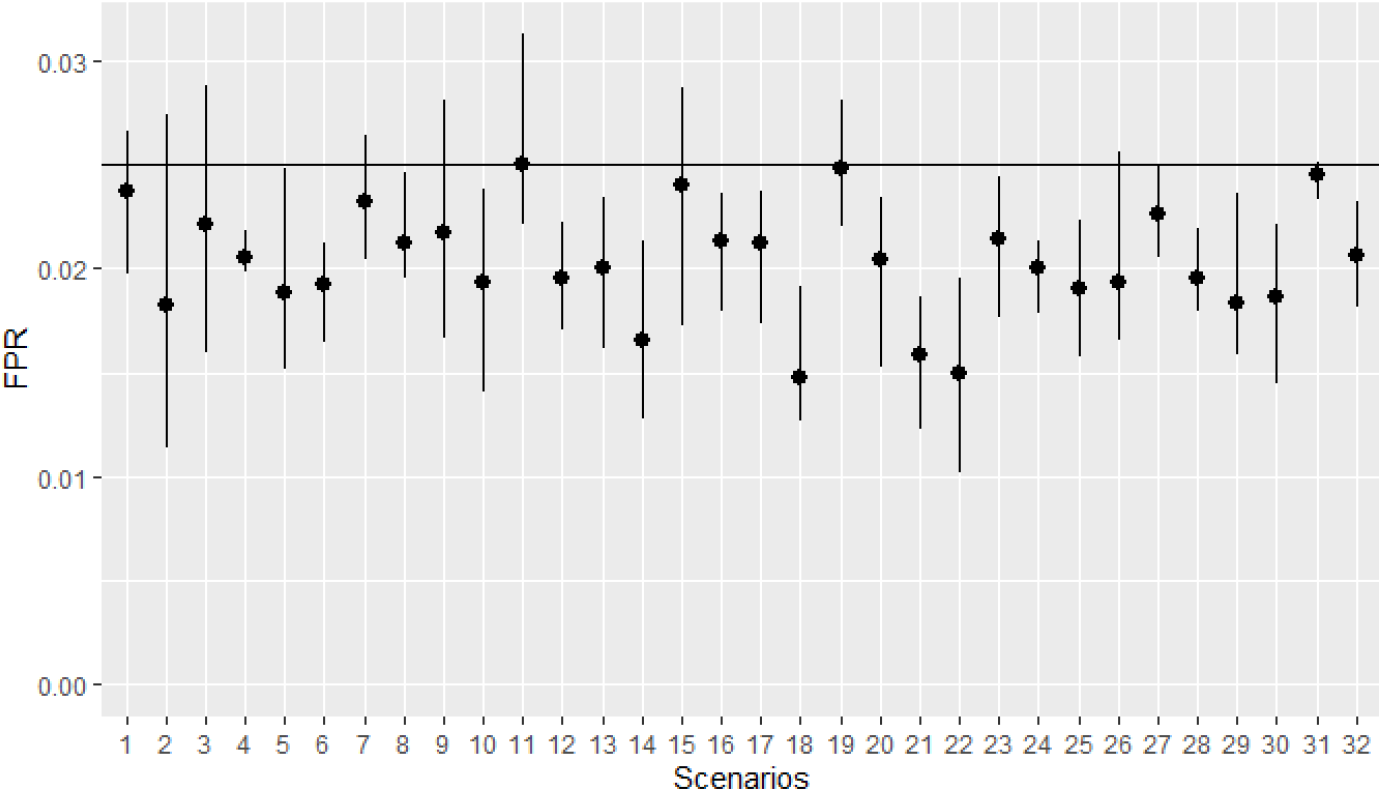
False positive rates obtained *α* = 0.05, *s* = 2.5 and *k* = 3. The horizontal line represents the nominal value of 0.025 and the numbers refer to the scenarios.

### 4.2 Probability of detection

Figure 2 shows the PODs obtained for *α* = 0.05, *s* = 2.5 and *k* = 3. The point represents the median POD across the 5 iterations and the vertical line represents the range of PODs for those 5 iterations. PODs are varying around 0.5 with higher value for scenarios with covariates.

**Figure 2:**
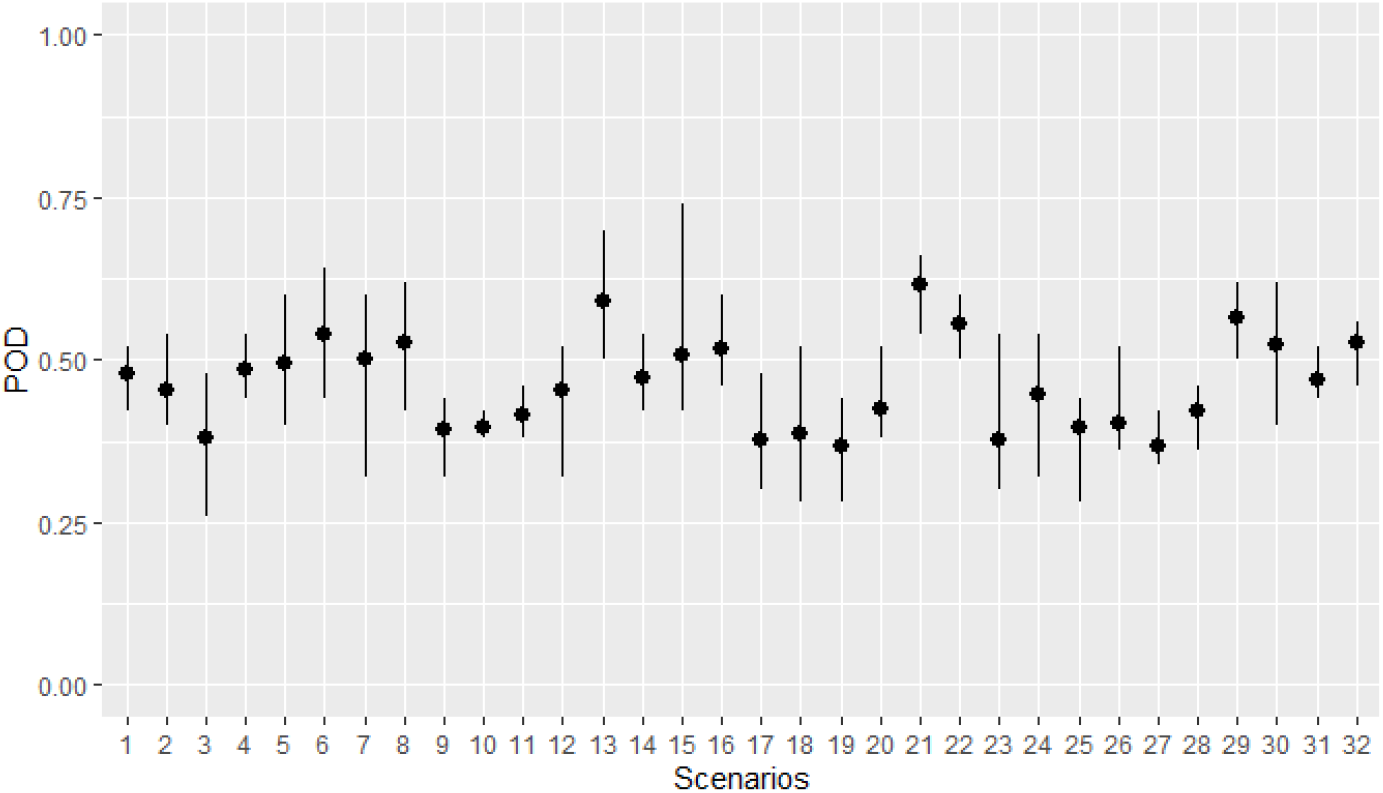
Probability of Detection obtained *α* = 0.95, *s* = 2.5 and *k* = 3. Numbers refer to scenario.

### 4.3 Exploratory analyses

The previous simulations fixed some parameters that can have an impact on the results of the model as *s* or *k*. To check the impact of those parameters, we performed some quick exploratory analyses. using the 7th scenario which includes every parameter with a medium value.

#### Weight threshold

Figure 3 shows the FPRs and the PODs obtained for different weight thresholds *s* = 1, 1.5, 2, 2.5 and 3, for *k* = 3 and *α* = 0.05.

**Figure 3:**
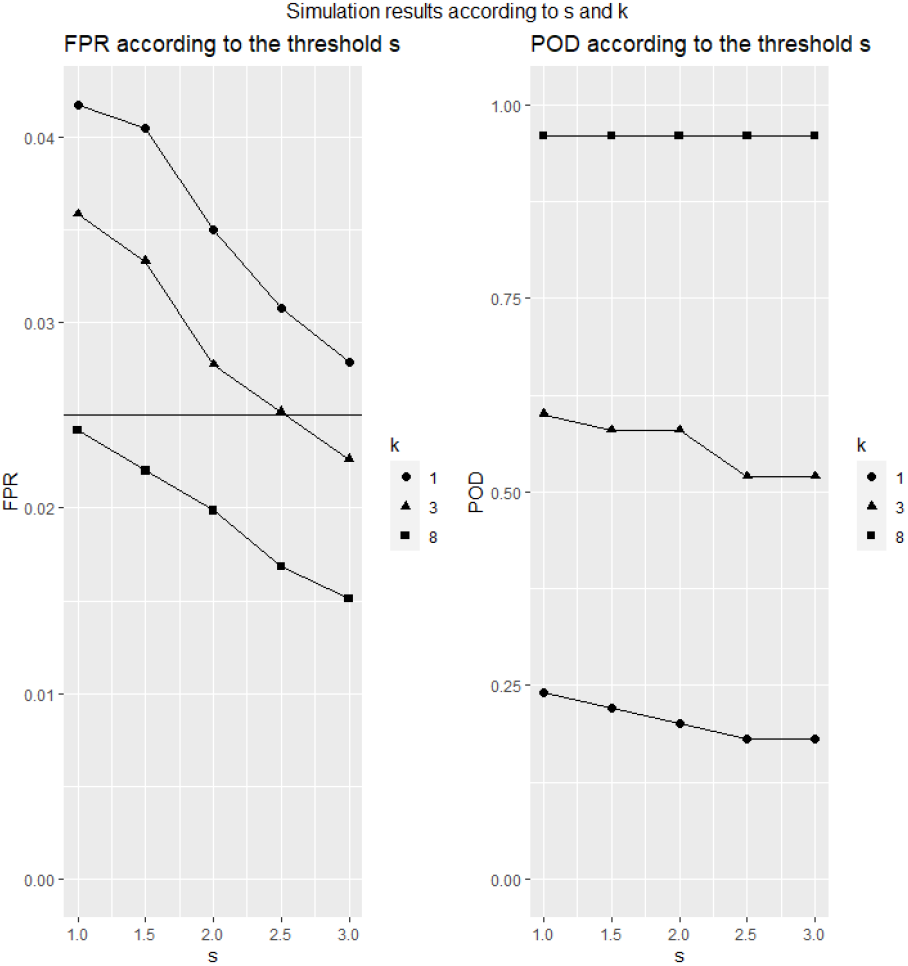
False Positive Rate and Probability of Detection according to the threshold s for outbreak sizes k=1, 3 and 8.

As expected, we see that a higher threshold *s* leads to lower FPR: the underweighting is less strict which leads to less alert. The optimal *s*, which is the *s* that gives a FPR around the nominal value, is different according to the size of the outbreaks of our dataset. For large outbreaks (*k* = 8), we should choose a low *s* around 1. For medium outbreaks (*k* = 3), we should choose a *s* around 2.5 (which is our baseline value for our previous simulations). For small outbreaks (*k* = 1), a higher *s* should be chosen (*s* = 3). Those results are quite consistent with the results from Noufaily *et al*. (2013) [11] that found that the optimal value was *s* = 2.58.

The POD remains almost constant for all values of *s*: the choice of the threshold *s* will mostly influence the FPR and is used to monitor the specificity of the model.

#### Outbreak size

Figure 4 shows the FPRs and the PODs obtained for different outbreak size *k* (from 1 to 10) for *s* = 2.5 and *α* = 0.95. The same baseline dataset is used for the different *k* (only the outbreaks are modified) to isolate the impact of the outbreak size.

**Figure 4:**
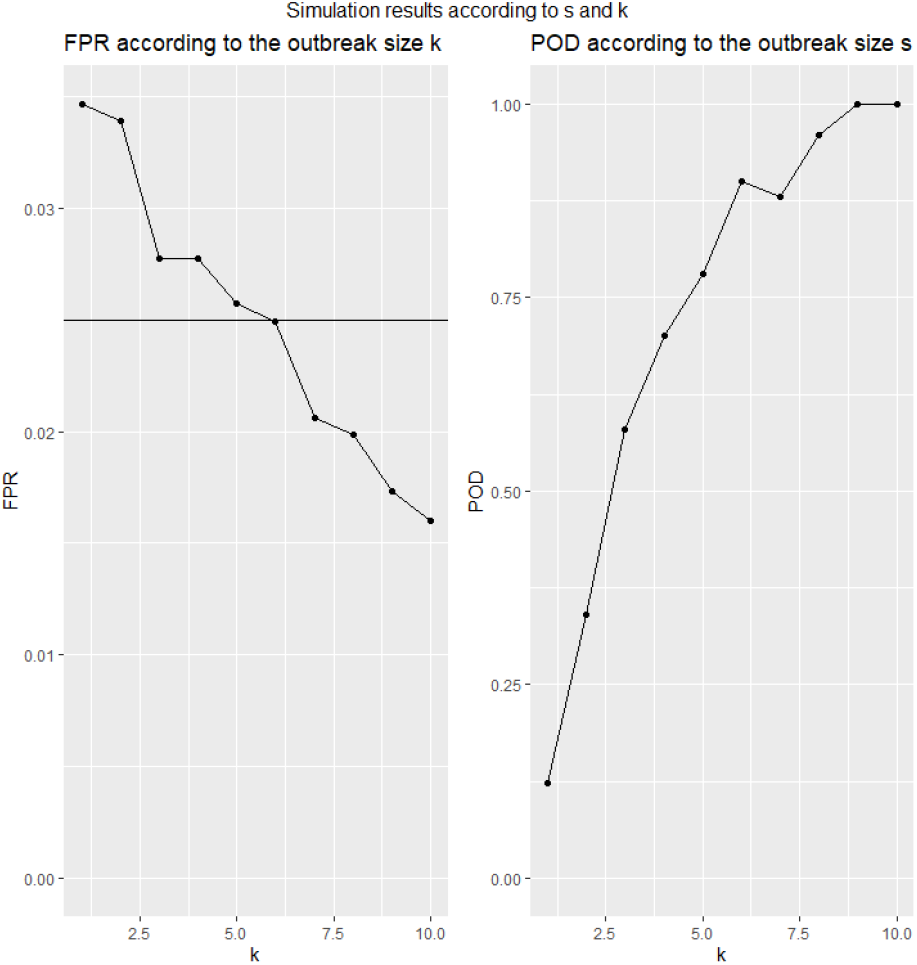
False Positive Rate and Probability of Detection according to the outbreak size *k* for *α* = 0.05 and *s* = 2.5

The FPR is lower when the outbreak size is higher: it underlines yet again that the *s* should be adjusted according to the expected size of the outbreaks.

On the other hand, the POD increases greatly with the value of k to approach almost 100% after *k* = 8.

## 5 Case study: sick leave monitoring and the example of Covid-19

### 5.1 Data

The dataset comes from digital files from companies insured by Malakoff Humanis, a French health insurer. The insured companies monthly updates the socio-demographic characteristics of their employees, their administrative status and their sick leaves. The dataset is named *Déclaration Sociale Nominative* and describes all employees of 1376 French companies which have more than 50 employees, followed since January 2018.

The outcome of the model is the weekly number of sick leave days for each company and. The number of theoretical work days per week is included in the model as an offset to adjust for the size of the company.

The dataset also describes some characteristics of the companies which we include in the model as covariates: the number of employees per category of age (35 years old and less, 36-45 years old, 46-55 years old, 56 years old and more), the number of workers with a temporary contract and the number of workers per occupational categories (managerial occupations, intermediate occupations, manual lower occupations, non-manual lower occupations). We also add an indicator of week with high numbers of vacation days that correspond to low level of sick leaves. A report of the statistics departement of the French Ministry of Labour (DARES) shows that peaks in annual leave occur during the Christmas school holidays (last week of December and first week of January) and during summer (second week of July to third week of August) [21].

We train the model on 2018 and 2019 and evaluate it on data from January to May 2020. We set the type I error to be *α* = 0.05 and the reweighting threshold to *s* = 2. We define an outbreak when we observe during two consecutive weeks a sick leave rate above the alert threshold.

### 5.2 Results

Figure 5 shows the evolution of the mean sick leave rate from January 2018 to May 2020. 2020 is a special year because of the COVID-19 pandemics. Before the third week of March 2020, the sick leave rate follows a distribution similar to the previous years. A peak is observed at the third week of March 2020 and corresponds to the first week of lockdown in France. This high sick leave rate should not be interpreted as a high incidence of COVID-19 patients but as an implication of regulatory change: employees who had to stay home for their children were provided sick leaves. We run the algorithm on 2020 data to identify companies which were impacted by COVID-19 after the lockdown and companies which had alerts non-related to COVID-19 before lockdown.

**Figure 5:**
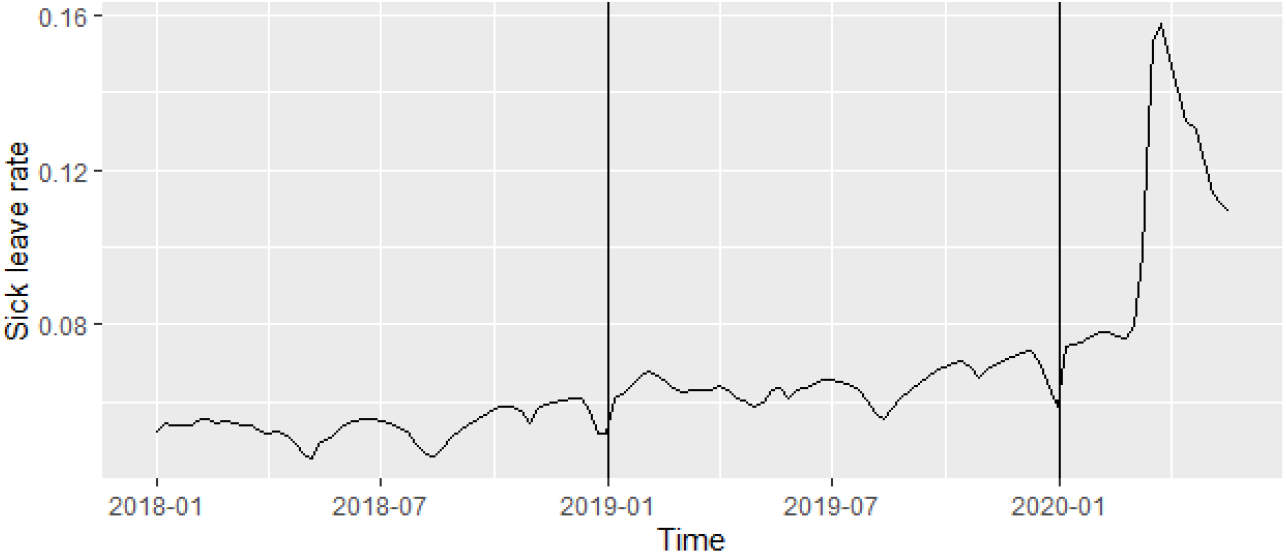
Weekly mean sick leave rate among all companies

Table 2 gives the results of the algorithm run on the dataset in 2020. We observe that, before lockdown, 5.9% and 7.9% of the companies have an outbreak in January and February. In March, the number of companies in outbreak rises to 56.8% in March, 58.9% in April and 42.1% in May. More than half of the companies therefore seem to have been affected by COVID-19 in terms of sick-leave. The companies with an outbreak have higher sick leave rate after lockdown (17.7% in April and May) than before (8.3% in January and February).

**Table 2:** Proportion of companies with a declared outbreak in the five first months of 2020

|  | January | February | March | April | May |
| --- | --- | --- | --- | --- | --- |
| Number of companies with an outbreak | 5.9% | 7.9% | 56.8% | 58.9% | 42.1% |
| Sick leave rate among companies with outbreak | 7.5% | 7.6% | 9.6% | 8.0% | 6.5% |
| Sick leave rate among companies with no outbreak | 8.3% | 8.3% | 14.4% | 17.7% | 17.7% |

Figure 6 shows four examples of companies sick leave rates with different numbers of employees and outbreak incidences. The first company represents a case where an outbreak occurs just after the lockdown and then the sick leave level goes back to the baseline level. The second company presents no outbreak. The third company has a large outbreak just after the lockdown and the sick leave level stays really high. The fourth company has an outbreak at the beginning of the year. We can observe that the alert threshold is consistent with the baseline level of absence of each company.

**Figure 6:**
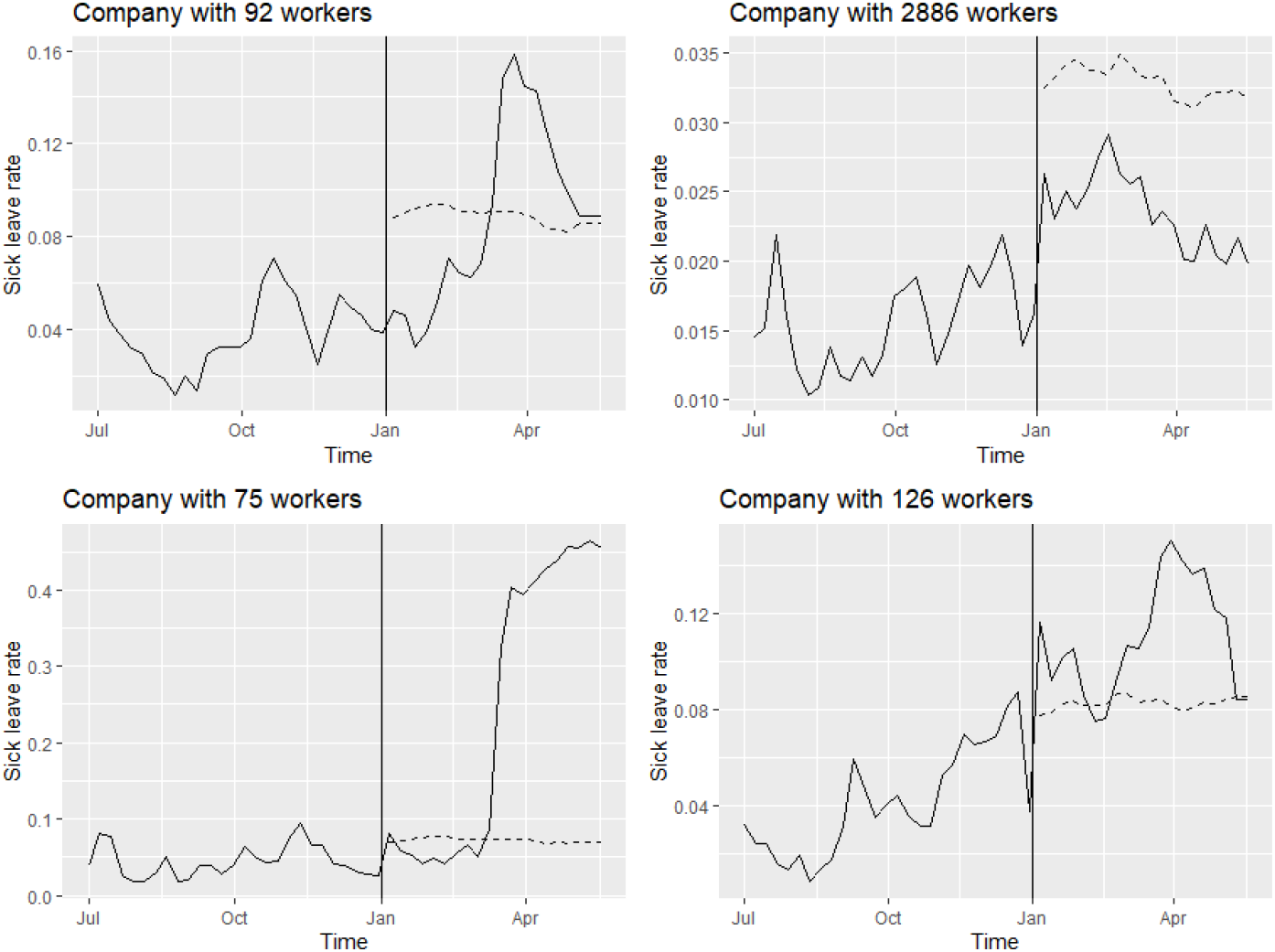
Four examples of companies sick leave rates from July 2019 to May 2020. The vertical line represents the first day of 2020, the solid line represents the weekly sick leave rate of the company and the dashed line represent the alert threshold.

## 6 Discussion

We proposed an adaptation of the Farrinton algorithm for surveillance of multisite data and we proposed an application to the case of sick leave. The inclusion of a random effect resulted in change in the choice of the weight function and of the alert threshold. Extensive simulations proved that the model provides results that are consistent with the results of the flexible Farrington algorithm. We found that the optimal *s* was 2.5 and it led to average FPR between 0.015 and 0.025 and POD between 0.368 and 0.616 according to 32 different scenarios and for an outbreak size of 3. As expected, higher *k* led to higher POD and lower FPR.

Computation time could be an obstacle for mixed effect models as mentioned [12]. It is still an issue with this algorithm as approximately an hour and a half was needed to run an iteration of the model for 1376 companies on our computer. To improve this computational weight, the model could have been stratified by group of companies (by company size for instance) and the new models could have been run in parallel. Some recent developments show the interest of neural networks for the estimation of generalized linear mixed models: this could be another alternative to improve the performance of our model [22]. These difficulties lead us to think carefully about the need to use a mixed model: this model is not necessarily the most appropriate one for each situation.

The application to sick leave provides interesting results in the case of COVID-19 and helped to indentify companies that were impacted by the pandemics. We did not use 5 years of historical data as suggested by Noufaily *et al*. (2013) [11] because our data did not allow us to do so. However, we believe that this model can be fitted on a smaller time window since this is compensated by a large number of companies.

In more standard situations, this surveillance system could help to identify and alert companies that have unusual levels of sick leave in a timely manner to monitor potential issues as bad management practices. Mixed model surveillance is already used in practice to monitor some syndromic data [12] and this study provides a validated algorithm including reweighting procedure.

## Data Availability

The data that support the findings of this study are available from the corresponding author, Tom Duchemin, upon reasonable request.

## Acknowledgments

The first author acknowledges Association Nationale de la Recherche et de la Technologie (grant number 2017/1517).

## Supporting information

R codes used for the analyses and the simulations can be found at: https://github.com/TomDuchemin/mixed_surveillance.

